# Cutaneous leishmaniasis in Casablanca-Settat region (Morocco): spatio-temporal analysis of disease dynamic and machine learning based case prediction

**DOI:** 10.1101/2025.01.28.25321261

**Authors:** Imane El Idrissi Saik, Hasnaa Talimi, Bouchaib Badri, Souad Bouhout, Rachida Fissoune, Myriam Riyad, Meryem Lemrani

## Abstract

Cutaneous leishmaniasis (CL) caused by *Leishmania* protozoa and transmitted through infected sandfly bites, poses a significant public health burden in Morocco. Our research aims to retrospectively assess spatio-temporal patterns of CL in the most densely populated region of the country, Casablanca-Settat, during a period of 14 years (2009–2022). We investigate epidemiological trends, seasonal fluctuations and climatic factors influence on CL prevalence and predict cases in the region’s most important CL focus. Using Geographic Information Systems (GIS) and statistical methods, we analyzed CL data obtained from the Moroccan Ministry of Health and Social Protection with demographic information and climate-related parameters. Four machine learning prediction models for CL cases in the El Brouj focus were compared and their performance was evaluated. Our results revealed a steady increase in CL cases until 2019, followed by a significant surge, with *Leishmania tropica* identified as the causative agent for all documented cases. The incidence rates varied among demographic groups, with higher rates observed in females and children aged 5-14. Seasonal analysis indicated a notable increase in cases during spring. Spatial distribution analysis identified El Brouj as the sole endemic CL focus_and the highest hotspot in the area, underscoring the necessity of targeted interventions in this location. Furthermore, Spearman correlation analysis revealed a significant association between CL cases and low temperature and precipitation, emphasizing climate variables role in disease incidence. Prediction modeling for future CL cases in El Brouj focus was assessed with six different models and Linear Regression was the most suitable. This retrospective study provides crucial insights into CL epidemiology in the Casablanca-Settat region, highlighting the need for implementing a coordinated approach tailored to the region that could help curb the spread of CL in Settat province, given the predictive model’s outlook on future CL trends in this area.

## Introduction

Cutaneous leishmaniasis (CL) is a neglected tropical disease caused by a flagellated protozoan belonging to the *Leishmania* genus. It is transmitted through the bite of an infected sand fly and the disease encompasses a wide range of symptoms going from mild cutaneous lesions to severe ones [1]. Over 20 species of *Leishmania* parasites contribute to diverse clinical symptoms in both humans and animals. According to the World Health Organization (WHO), CL is endemic in 92 countries and more than one billion individuals are at risk of contracting the illness [2]. CL does not only affect humans but is rather a One Health issue, as sylvatic mammals serve as primary hosts such as dogs and rodents who act as reservoirs for many *Leishmania* species [3]. CL is characterized by a variety of clinical presentations that may be due to many factors, mainly the infecting *Leishmania* species and the individual’s immune response. In some cases, the lesions may heal spontaneously, while in others, they can persist and lead to disfiguring scars. Thus, CL is a disease that causes a significant psychological burden as patients are often stigmatized, particularly those living in low socio-economic settings [4]. Moreover, the treatment of CL is heavy and has many undesirable side effects, especially intramuscular meglumine antimoniate (Glucantime™) [5].

The Kingdom of Morocco is endemic to CL caused by three *Leishmania* species. *Leishmania major* infection results in the zoonotic cutaneous form mainly in the southeastern region [6]. While responsible for visceral leishmaniasis, *L. infantum* used to cause sporadic cutaneous cases, but it is now shifting towards endemicity in foci such as Taounate, Taza and Sidi Kacem [7] and more recently, the city of Salé (Rabat-Salé-Kénitra region), according to the Department of Parasitic Diseases at the Ministry of Health. In addition, *L*. *tropica,* a species transmitted through the bite of female *Phlebotomus sergenti,* became the most widespread geographically [8], and even though animal infections were reported, it is still mostly considered anthroponotic [9].

Anticipated changes in climate are expected to broaden both the geographical reach and activity periods of phlebotomine vectors. In Morocco, the elevation in minimum temperatures has bolstered the survival of sand fly larvae throughout winter, extending their activity period. Temperature has a significant effect on the *Leishmania* development cycle within its vector, as well as the sand fly’s metabolism and developmental period [10–12]. This, in conjunction with increased human movements, is believed to be a significant factor contributing to the dissemination of CL. Therefore, it is essential to investigate the spatio-temporal trends of the disease nationwide, particularly in areas with active CL foci, in order to aid in the implementation of effective preventive measures.

Casablanca-Settat is the most densely populated region of the territory [13]. It is located in the center-west bordering the Atlantic coast. In this region, a specific focus of CL emerged in 2006 in the Settat Province, which had previously not been affected by CL. Since then, CL due to *L. tropica* has become endemic in this province, and from 2007 to 2012, approximately 553 cases were reported from El Brouj, a locality in Settat province [14]. The present manuscript will discuss the cases reported between 2009 and 2022 from El Brouj, Settat Province along with a more comprehensive analysis of any significant trends or outbreaks observed during this period. As no reservoir has been identified for *L. tropica* in Morocco yet, the disease cycle mainly relies on the spread of *P. sergenti* through human movements. Considering the wide geographical distribution of *P. sergenti,* the increase in population density, population movements, climate change, and the spread of CL due to *L. tropica* in new foci previously unknow, there is an urgent need for the assessment of spatio-temporal trends of CL in Casablanca-Settat region [9]. Indeed, a number of environmental factors amongst which climate variables are associated with high CL incidence. [15]

Machine learning (ML) application to CL case prediction has gained momentum in recent years as they can efficiently analyze large amounts of epidemiological data. Disease detection and classification is improved through the use of ML models, while also allowing for epidemic forecasts [16]. Therefore, our present retrospective study aims to characterize the epidemiological trends in the incidence of CL in the Casablanca-Settat region over a 14 years period between 2009 and 2022. Moreover, we study the association of climate variables with the disease incidence in the region and we identify the spatial patterns of the disease using Geographic Information Systems (GIS), and assess the most suitable ML prediction model to predict CL cases in El Brouj, the only focus identified in the region.

## Material and methods

### Study area

Located in the center-west of Morocco **(Fig 1)**, the region of Casablanca-Settat harbors the economic capital and is the most densely populated region of the country. It is located at 150 m above the sea level and according to a 2014 Moroccan official census, the population reached 6,862 million people, living at a density of 353 people per km^2^ and the region extends on 19,448 km^2^ [17]

**Fig 1:**
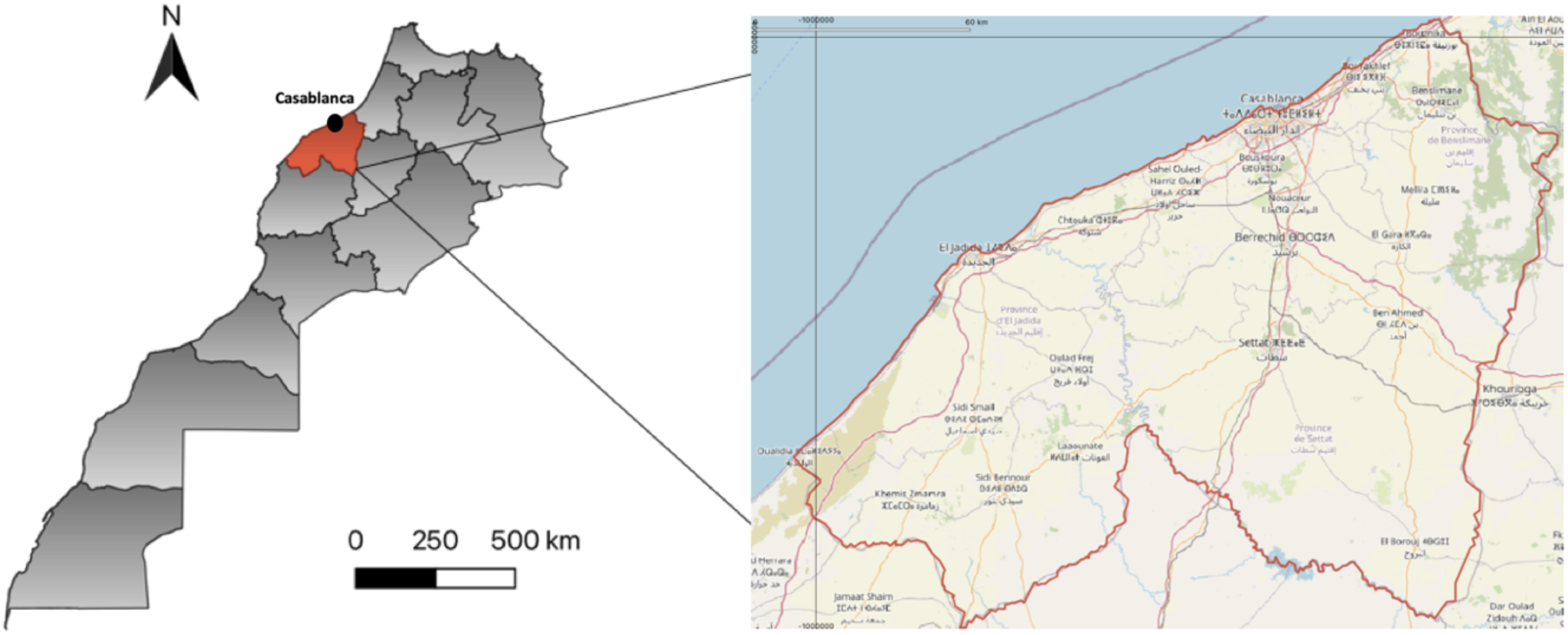
Geographical location of the study area: Casablanca-Settat region *(made with QGIS3.8),* base map and data from OpenStreetMap and OpenStreetMap Foundation.

### Ethics Statement

The approval for this study was obtained from the Ministry of Health and Social Welfare in Morocco. Since data were collected during routine active surveillance, no parents were interviewed and no patients’ names were recorded. Thus, ethical approval was not required for this study. Fully anonymized data were accessed on November 21st 2023.

### Data collection

Epidemiological Data were collected in Excel format from the Directorate of Epidemiology and Diseases Control (Direction de l’Epidémiologie et de la Lutte contre les Maladies, DELM), at the Moroccan Ministry of Health and Social Protection. The spreadsheets contained 712 cases of CL cases documented in Casablanca-Settat region throughout the 14 years period extending from 2009-2022. The variables age, sex, date of disease onset, origin, *Leishmania* genotyping result and patient location were also provided.

Demographical data were obtained from the High Commission of Planning (Haut-Commissariat au Plan, HCP): https://www.hcp.ma/reg-casablanca/Direction-Regionale-de-Casablanca-Settat_r7.html

Climatic data was extracted from the NASA’s POWER database available at https://power.larc.nasa.gov/data-access-viewer/. Monthly and annual data of Mean/Minimum/Maximum Temperature, Precipitation and Relative humidity were obtained for the El Brouj locality for the period from 2009 to 2022.

### Data Mapping

The geographical distribution of CL was mapped using free QGIS 3.28 software. The shapefiles of regional boundaries were obtained from the DELM. All patient locations provided by the Ministry of Health were geocoded manually using Google’s My Maps software (https://www.google.com/maps/about/mymaps).

### Statistical analysis

Annual incidence-rates and their confidence interval (95%) were calculated by sex, age, infecting species and whether the infection was autochthonous or not. Therefore, the research period was split into three periods in order to test for temporal variations in the effect of these factors on the incidence of leishmaniasis: 2009–2013 (n=140), 2014-2018 (n=137), 2019-2022 (n=435). These time intervals allow for a clear comparison of changes in the incidence of the disease over time. Statistical analyses were conducted using R version 4.2.2. Our R analysis was structured into three main scripts, each addressing specific aspects of the data.

First script focused on calculating the incidence rates and their confidence intervals. For this, we utilized several R packages such as purrr, dplyr, gt, readxl, and epiR. The raw data was imported from an Excel file using read_xlsx. We then defined a function, calculate_incidence, to compute incidence rates alongside their 95% confidence intervals, expressed per 100,000 individuals. The dplyr package facilitated data manipulation, enabling us to append the calculated incidence rates and confidence intervals to our dataset.

Secondly, a script was dedicated to visual data exploration, particularly examining the seasonal variation of cases. Here, ggplot2 was employed for its robust graphical capabilities. The script generated a line graph illustrating the number of cases across different seasons and periods, with distinct color codings for each period. This visual representation was crucial for identifying temporal trends and patterns in the data.

To reveal the temporal dynamics and seasonal fluctuations in the incidence of CL, we applied a generalized additive model (GAM) using “mgcv” package in R, which is capable of handling both linear and non-linear relationships while remaining interpretable. The response variable was the number of cases, while explanatory variables included period (2009-2013; 2014-2018; 2019-2022) and season (spring, summer, autumn and winter). The reference categories selected were the period 2014-2018 and Autumn.

Before performing a correlation test, we assessed the linearity between the variables (mean temperature, maximum temperature, minimum temperature, precipitation, and relative humidity) and the number of cases using the Shapiro-Wilk test with the package stats in R to evaluate the normality of the data distribution, a key assumption for linear relationships.

The fourth script examined correlations between environmental factors and the number of cases. We created scatter plots with regression lines to show the relationship between each factor (e.g., temperature, humidity, precipitation) and case counts. Spearman’s rank correlation was used to measure the strength of these relationships, with correlation coefficients and p-values added to the plots. The cor.test() function from the stats package was used to perform the correlation tests. The plots were arranged in a grid for easy comparison.

### Prediction Methodology

#### Dataset

The dataset on which the analysis is based includes a comprehensive compilation of CL cases recorded each year between 2009 and 2022. These cases are accompanied by simultaneous environmental and meteorological measurements, including temperature, relative humidity and precipitation, all of which significantly influence the disease’s transmission vectors and life cycle. The breadth of the data set makes it possible to examine long-term trends and assess the temporal consistency of the relationship between disease incidence and environmental factors.

#### Methodology

To predict the incidence of CL, we adopted a strong methodological approach utilizing four distinct supervised machine learning models. These models were trained on data spanning from 2009 to 2018, including both CL case counts and corresponding climate data, to generate forecasts for the years 2019 to 2022. The model that most closely reflected the actual data recorded during these years was then selected for future predictions for the next ten years, from 2022 to 2032.

#### Supervised Machine Learning Models for Predicting CL Incidence

The six supervised machine learning models used to predict CL incidence are Random Forest, Linear Regression, Gradient Boosting Regressor (GBR), Extreme Gradient Boosting (XGBoost), Decision Tree Regressor, and Support Vector Regressor (SVR). Each model was carefully selected to capitalize on its strengths and address the specific characteristics of the data:

##### Random Forest

This model excels in handling nonlinear relationships and interaction effects, making it well-suited for capturing complex patterns within the dataset.

##### Linear Regression

Implemented as a baseline model, it facilitates a straightforward interpretation of data relationships through linear associations.

##### Gradient Boosting Regressor (GBR)

Known for its robustness in modeling non-linearities and interactions, GBR was used to enhance predictions that involve complex data structures.

##### Extreme Gradient Boosting (XGBoost)

Selected for its advanced capability to model nonlinear boundaries using gradient boosting frameworks, XGBoost is particularly effective in dealing with diverse and intricate data distributions.

##### Decision tree regressor

simple and capable of handling both linear and non-linear relationships, making it useful for understanding complex data structures.

##### Support vector regressor (SVR)

famous for its ability to find a hyperplane that best fits higher-dimensional data, it generally handles complex relationships well.

The predictive outcomes are illustrated in six distinct graphs, each depicting the model’s projections for future CL cases. These visualizations vary in complexity and are based on different assumptions about future trends, thus providing a spectrum of perspectives on the potential progression of the disease.

#### Evaluation Parameters

Model evaluation was based on a series of performance measures, including Mean Absolute Error (MAE), Mean Square Error (MSE), Root Mean Square Error (RMSE) and R², allowing us to assess and compare the accuracy and explanatory power of each model [18]. Predictive performance was visualized in graphs contrasting historical data with predicted trends, providing a clear representation of the alignment of each model with observed data. This multi-faceted predictive approach was designed not only to predict the future incidence of CL, but also to interpret the underlying epidemiological and environmental interactions encapsulated in the historical data.

Each of these metrics offers a unique perspective on the model’s predictive capabilities:

##### Mean Absolute Error (MAE)

Quantifies the average magnitude of errors in a set of predictions, without considering their direction. It’s the mean outcome of the absolute differences between predicted and actual values, providing a straightforward measure of prediction accuracy. It was calculated as follows:

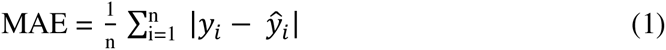

Where *y*_1_ is the actual value, 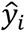 is the predicted value, and n is the number of observations.

##### Mean Square Error (MSE)

Measures the average of the squares of the errors, effectively giving greater weight to larger errors [18]. It’s used to assess the quality of a predictor by indicating how close the predicted values are to the actual values. It was calculated as follows:

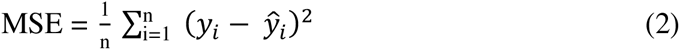

##### Root Mean Square Error (RMSE)

It is the square root of MSE and similarly evaluates the magnitude of the errors in predictions [18]. By square-rooting the MSE, RMSE converts the error metric back to the same unit as the target variable, making interpretation more intuitive. It was calculated as follows:

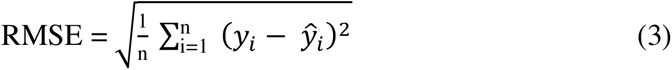

##### R-squared value (R^2^)

also known as the coefficient of determination, indicates the proportion of variance in the dependent variable that is predictable from the independent variables [18]. It provides a measure of how well the observed outcomes are replicated by the model. It was calculated as follows:

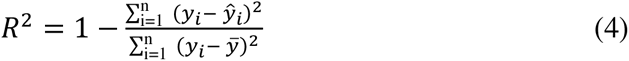

Where *ȳ* is the mean of the actual values.

Each of these parameters serves a specific purpose in model evaluation, providing distinct insights into predictive performance.

## Results

### Temporal variations of CL

During the study period (2009-2022), 712 cases of CL were notified to the Ministry of Health and Social Protection in the Casablanca-Settat region **(Table 1)**. The only reported species was *L. tropica* and 95% of all cases were autochthonous to the region. During the two first periods from 2009 to 2018, CL incidence was steady, with a slight decrease during the period 2014– 2018. However, from 2019 to 2022, an increase in CL incidence was observed, with more than four times the number of recorded infections in previous time frames. The cumulative incidence during the study period was 9.61 cases per 100,000 inhabitants.

**Table 1:** Incidence of leishmaniasis per 100.000 inhabitants and confidence interval 95% by complete study time and period 2009-2013 (n=140), 2014-2018 (n=137), 2019-2022 (n=435) in the Casablanca-Settat region.

|  |  | 2009-2013 | 2014-2018 | 2019-2022 | Total |
| --- | --- | --- | --- | --- | --- |
| **Sex** |  |  |  |  |  |
|  | Female (n= 413) | 1,12 [0,89-1,39] | 1,10 [0,88-1,37] | 3,75 [3,32-4,22] | 5,98 [5,43-6,56] |
|  | Male (n= 269) | 0,77 [0,58-0,99] | 0,74 [0,55-0,96] | 2,11 [1,80-2,47] | 3,63 [3,21-4,09] |
| **Age** |  |  |  |  |  |
|  | 0-4 (n= 72) | 0,17 [0,09-0,30] | 0,26 [0,16-0,41] | 0,52 [0,37-0,71] | 0,97 [0,76-1,22] |
|  | 5-14 (n= 227) | 0,45 [0,31-0,64] | 0,59 [0,43-0,79] | 2,01 [1,70-2,36] | 3,06 [2,67-3,48] |
|  | 15-24 (n= 78) | 0,18 [0,10-0,31] | 0,28 [0,17-0,43] | 0,58 [0,42-0,78] | 1,05 [0,83-1,31] |
|  | 25-44 (n= 144) | 0,47 [0,32-0,65] | 0,24 [0,14-0,38] | 1,22 [0,98-1,50] | 1,94 [1,63-2,28] |
|  | 45-64 (n= 170) | 0,54 [0,38-0,73] | 0,37 [0,25-0,54] | 1,37 [1,22-1,67] | 2,29 [1,96,6,56] |
|  | 65 or + (n= 21) | 0,05 [0,01-0,13] | 0,08 [0,02-0,17] | 0,14 [0,07-0,26] | 0,28 [0,17-0,43] |
| **Origin** |  |  |  |  |  |
|  | Autochthonous (n= 709) | 1,87 [1,57-2,21] | 1,82 [1,52-2,15] | 5,87 [5,33-6,45] | 9,57 [8,87-10,30] |
|  | Imported (n= 3) | 0,013 [0,0003-0,07] | 0,02 [0,003-0,09] | 0,00 | 0,04 [0,008-0,11] |
| **Species** |  |  |  |  |  |
|  | *L. tropica* (n=712) | 1,89 [1,59-2,23] | 1,84 [1,55-2,15] | 5,87 [5,33-6,45] | 9,61 [8,91-10,34] |
|  | *L. major* (n= 0) | 0,00 | 0,00 | 0,00 | 0,00 |
|  | *L. infantum* (n= 0) | 0,00 | 0,00 | 0,00 | 0,00 |
|  | **Total (n= 712)** | 1,89 [1,59-2,23] | 1,84 [1,55-2,15] | 5,87 [5,33-6,45] | 9,61 [8,91-10,34] |

An increase of the incidence was observed in all age groups, especially between the first period and the third period with the higher incidence found in the 5–14 age group (3.06 per 100,000 inhabitants). In addition, CL incidence according to sex followed the same pattern in all time periods as women are reportedly more affected by the disease than men (5.98 vs 3.63 total cases per 100.000 inhabitants, respectively) **(Table 1).** This trend has been observed every year except 2010, where a slightly higher number of cases was recorded in male patients **(Fig 2)**. Moreover, 2021 was the year with the peak annual incidence of CL, when in contrast, 2015 had the lowest peak **(Fig 2).**

**Fig 2:**
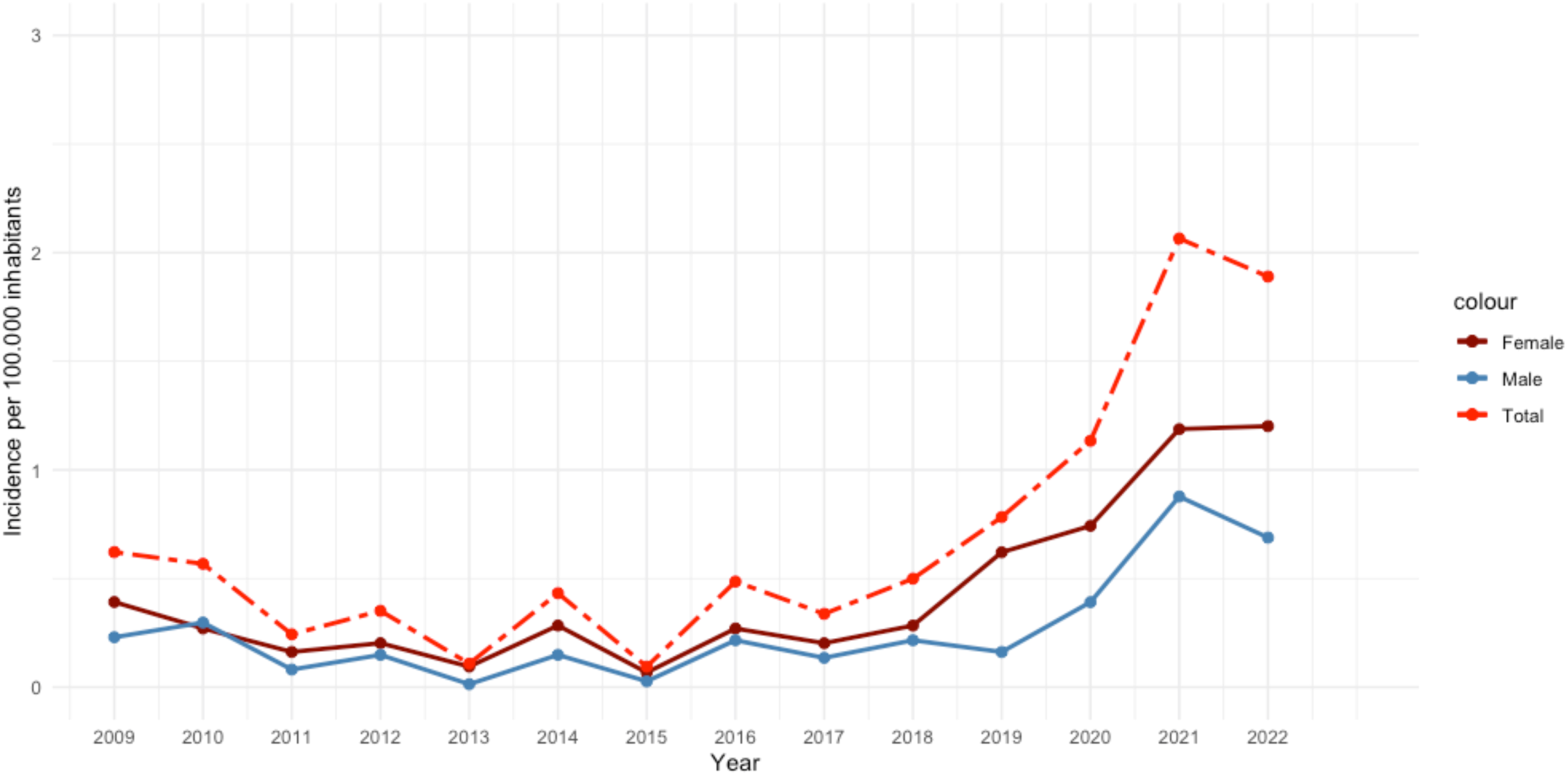
Temporal distribution of CL in Casablanca-Settat region: annual incidence by complete sample and sexes from 2009-2022.

### Seasonal variation of CL

We studied the variation of CL cases distribution depending on the season. **Figure 3** shows a seasonal trend of CL cases in Casablanca-Settat region, as spring (March-May) was the season with the highest number of CL cases recorded in the time periods 2009-2013 and 2019-2022. The highest number of cases ever recorded was 158 cases during the spring season of 2019-2022. During the 2014-2018 period, winter was the season with most cases (53 cases). Lower number of cases were recorded during summer and autumn of 2009-2013 period. Autumn was the season with the fewest cases overall.

**Fig 3:**
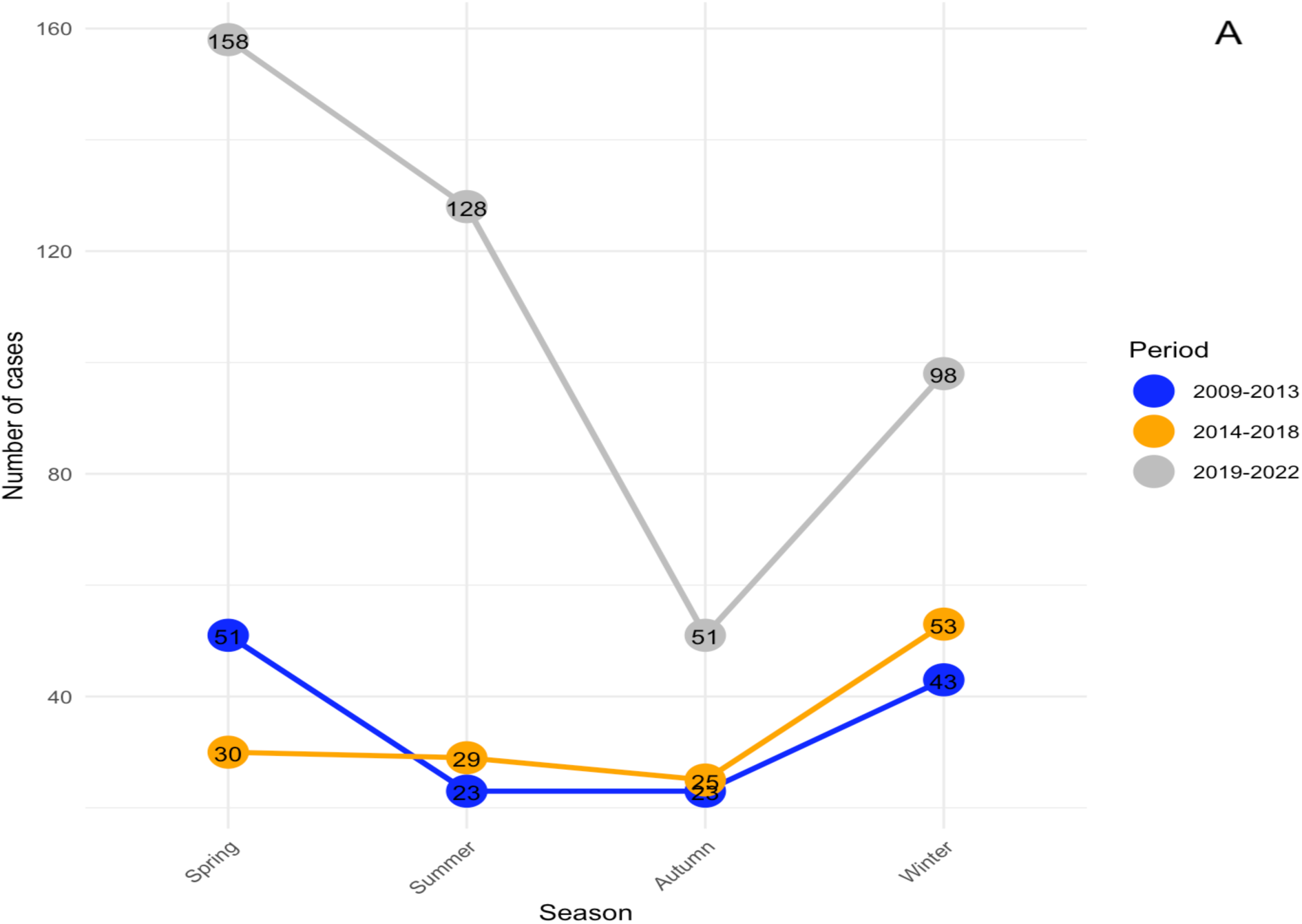
Seasonality of CL incidence from 2009-2022 in the Casablanca-Settat region

The parametric coefficients derived from the Generalized Additive Model (GAM) are presented in **Table 2**. The model suggests that Spring has a notable effect on CL incidence, with an estimated increase of 46.667 cases (p = 0.06697), although the result is only borderline significant. Similarly, Summer shows a positive, albeit non-significant, effect (Estimate = 27.000, p = 0.24387). In contrast, Winter presents a non-significant effect (Estimate = 31.657, p = 0.18046). A key finding from the model is the significant rise in CL incidence during the 2019-2022 period, with an estimated increase of 74.500 cases (p = 0.00624) compared to the reference period (2014-2018).

**Table 2:** Parametric coefficients of the GAM model.

| Term | Estimate | Std. Errot | T-value | P-value |
| --- | --- | --- | --- | --- |
| (Intercept) | 7.917 | 18.097 | 0.437 | 0.67709 |
| Season Summer | 27.000 | 20.897 | 1.292 | 0.24387 |
| Season Spring | 46.667 | 20.897 | 2.233 | 0.06697 |
| Season Winter | 31.667 | 20.897 | 1.515 | 0.18046 |
| Period 2009-2013 | 0.750 | 18.097 | 0.041 | 0.96829 |
| Period 2019-2022 | 74.500 | 18.097 | 4.117 | 0.00624 ** |

Overall, the GAM analysis supports the observation of heightened CL incidence in Spring during the 2019-2022 period, indicating a potential seasonal and temporal shift in disease occurrence in the Casablanca-Settat region.

### Spatial distribution of CL

The disease’s spatial distribution in the region shows that autochthonous cases are confined to the Settat province, with different geographic locations experiencing variable degrees of prevalence and discrete clusters **(Fig 4).** The highest incidence hotspot is clustered in El Brouj and in its nearby localities. El Brouj accounts for 96% of all recorded CL cases during the study time period (690 cases in total) and within this commune, cases are mainly present in the center. The prevalence is lower in other localities around the city of Settat harboring around 10 cases.

**Fig 4:**
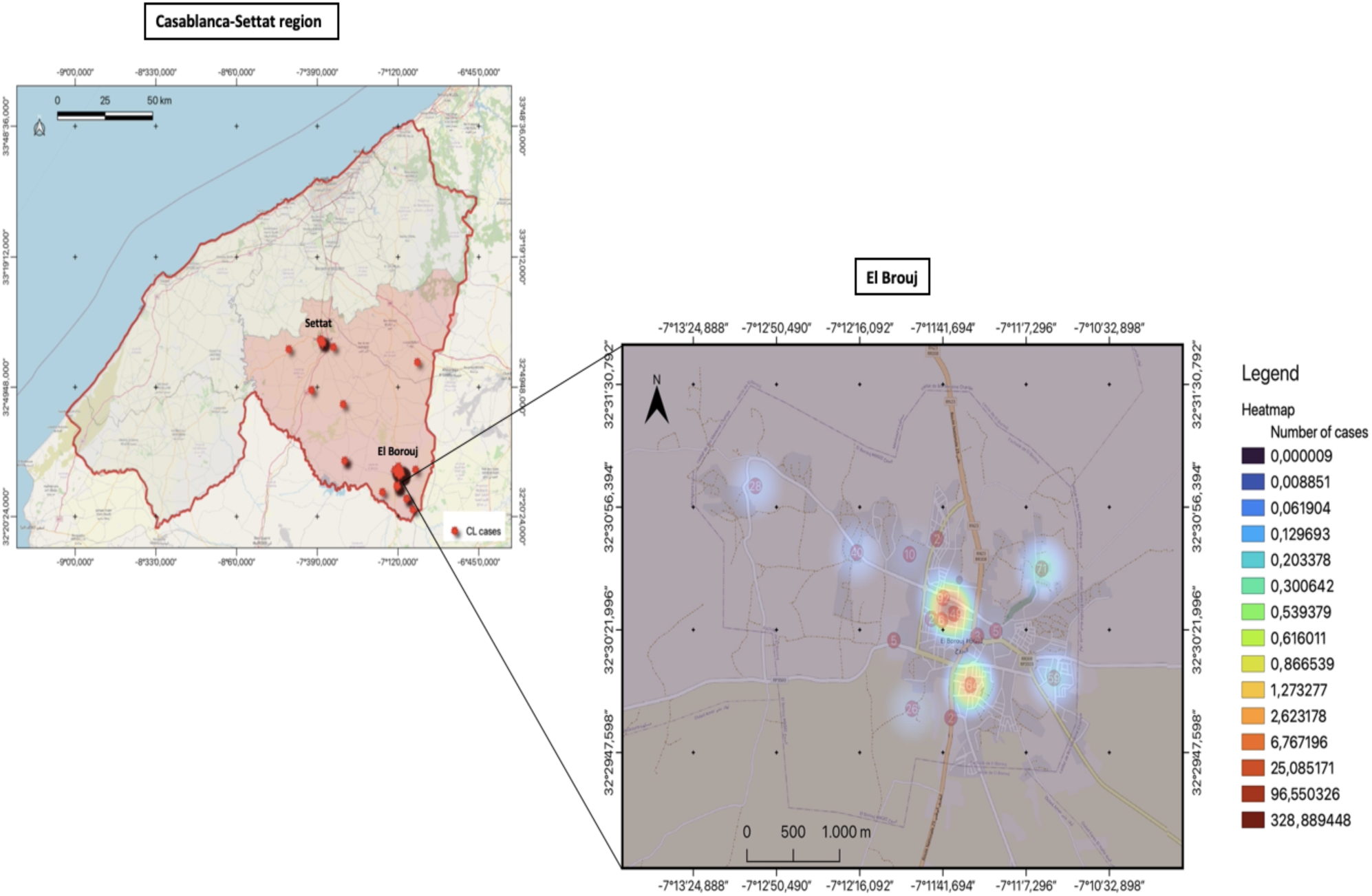
Heatmap of the spatial distribution of CL in the Casablanca-Settat region and El Brouj locality by complete study time 2009-2022. Figure made with *QGIS3.8,* base map and data from OpenStreetMap and OpenStreetMap Foundation.

The whole Casablanca-Settat region is divided into 9 provinces and no cases of CL were recorded during the study period in the other provinces of Sidi-Bennour, Berrechid, El Jadida, Casablanca, Nouaceur, Mediouna and Mohammedia.

### Spearman Correlation Analysis of CL cases with Climate Factors

In the context of understanding the impact of climatic factors on the distribution of CL in the Casablanca-Settat region, we have focused our study on the El Brouj area, which has the highest number of recorded cases. Prior to assessing the correlation between the number of cases in El Brouj area and climate variables, we performed Shapiro-Wilk test to assess linearity of variables’ distribution. Since not all p-values are < 0,05 **(Table 3),** we decided to use Spearman correlation test.

**Table 3:**
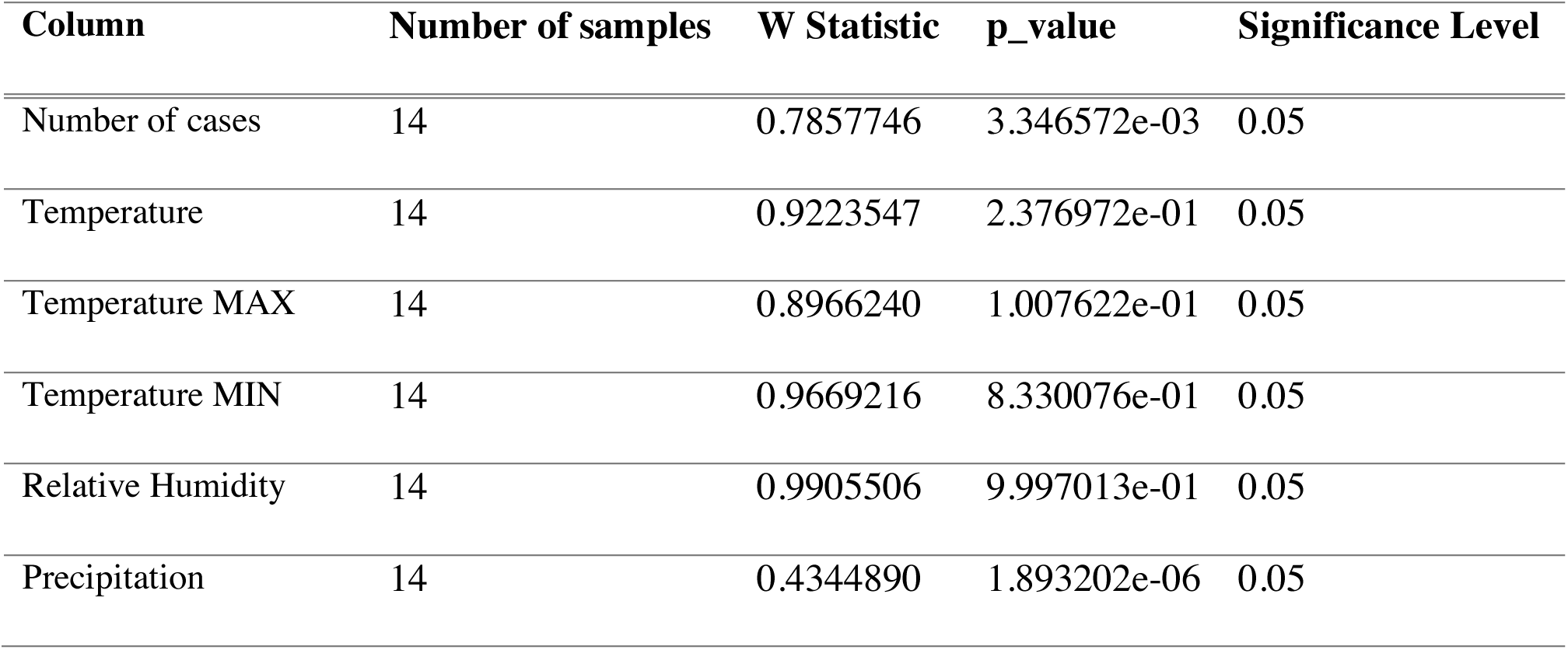
Shapiro-Wilk test results on the climatic variables and number of cases distribution. *Made with R version 4.2.2*.

| Column | Number of samples | W Statistic | p_value | Significance Level |
| --- | --- | --- | --- | --- |
| Number of cases | 14 | 0.7857746 | 3.346572e-03 | 0.05 |
| Temperature | 14 | 0.9223547 | 2.376972e-01 | 0.05 |
| Temperature MAX | 14 | 0.8966240 | 1.007622e-01 | 0.05 |
| Temperature MIN | 14 | 0.9669216 | 8.330076e-01 | 0.05 |
| Relative Humidity | 14 | 0.9905506 | 9.997013e-01 | 0.05 |
| Precipitation | 14 | 0.4344890 | 1.893202e-06 | 0.05 |

The scatter plots in **Figure 5** show the relationships between the incidence of CL cases and five different climatic factors (’Minimum Temperature’, ’Maximum Temperature’, ’Mean Temperature’, ’Precipitation’, and ‘Relative Humidity’), supplemented by linear regression lines. Each plot includes a shaded area representing the 95% confidence interval for the regression line.

**Fig 5:**
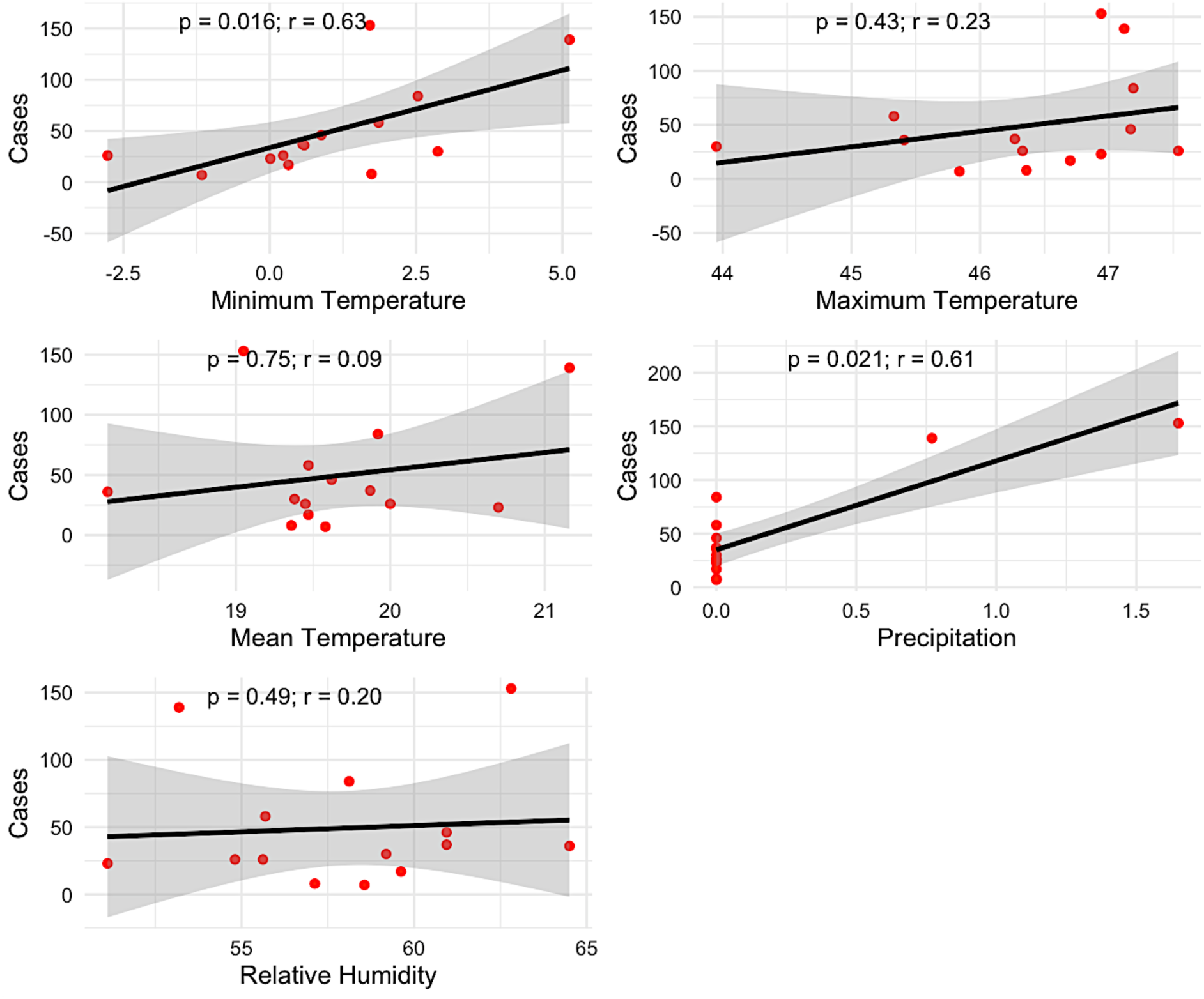
Spearman Correlation and Linear Regression of CL case numbers in El Brouj focus with climate data from 2009 to 2022.

Our results revealed a positive correlation between minimum temperature and cases, indicated by a correlation coefficient (r) of 0.63. The association is statistically significant, with a p-value of 0.016, suggesting that the observed correlation could potentially contribute to an increase in CL cases. Similarly, the plot for maximum temperature shows a positive but weak correlation (r = 0.23) with cases, but not statistically significant (P-value: 0.43). Conversely, mean temperatures were slightly positively associated with CL cases (P = 0.75, r = 0.09), suggesting that this association may rather be due to random variation. The most substantial correlation is observed with precipitation, showing a high correlation coefficient of r = 0.61, strongly statistically significant with a P-value of 0.021. This strong association suggests that increases in precipitation could be linked to a significant rise in the number of cases. However, the plot for relative humidity presents a negligible positive correlation with cases (r = 0.20) with no statistical significance (P = 0.49).

### Comparative analyses of machine learning models for predicting the incidence of CL

**Figure 6** and **Table 4** show the comparative findings of six different machine learning models that were used to forecast trends from 2009 to 2022. The results of various predictive models were displayed in each panel, showing historical data trends, corresponding predictions, and 95% confidence intervals.

**Fig 6:**
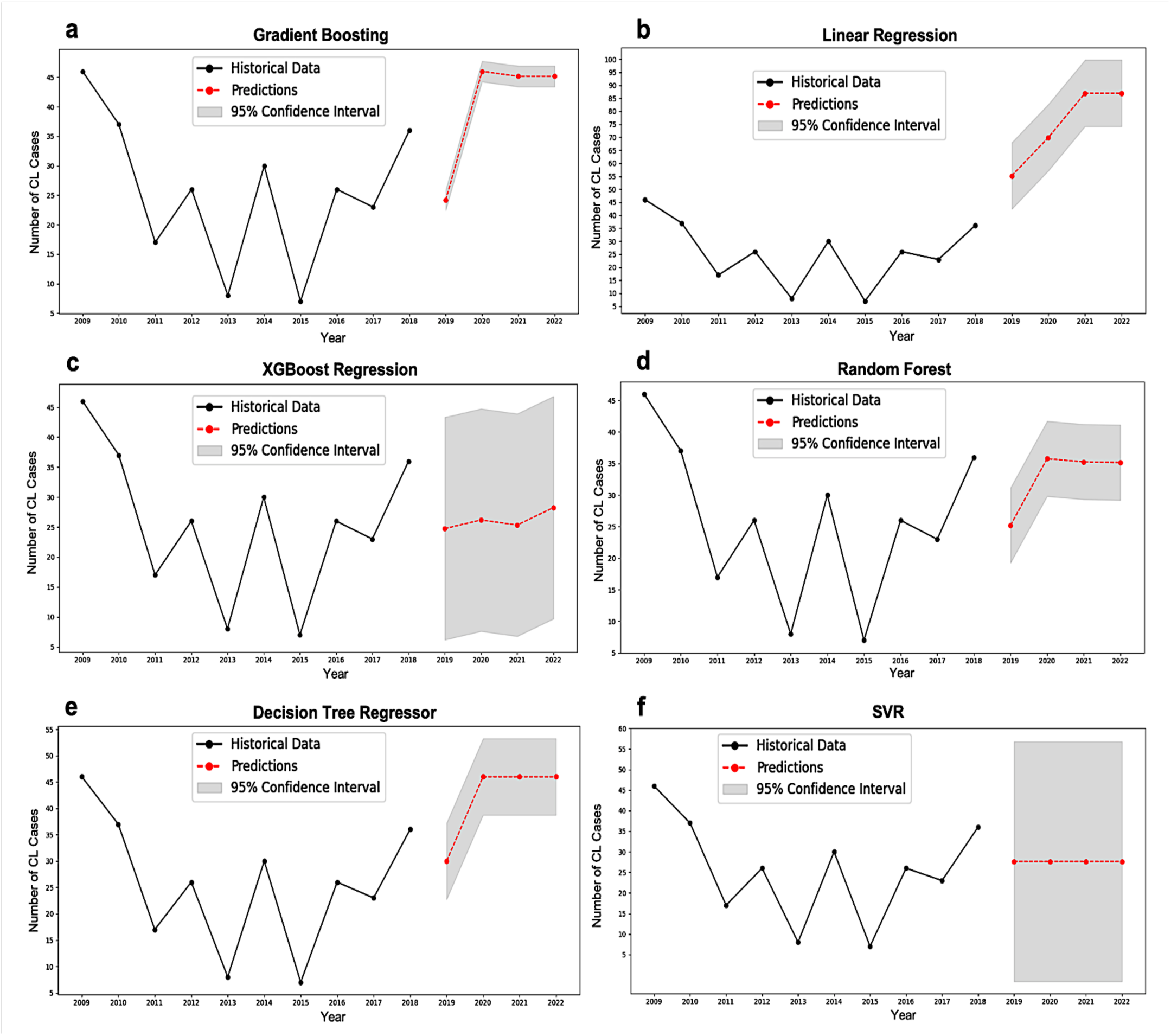
Predictive results from comparing six different machine learning models for CL incidence in El Brouj from 2019 to 2022 (95% confidence interval).

**Table 4:** Evaluation parameters of six different machine learning models for CL incidence.

| Models | MAE | MSE | RMSE | R<sup>2</sup> |
| --- | --- | --- | --- | --- |
| Gradient Boosting Regressor | 2.15 | 8.27 | 2.88 | 0.83 |
| Linear Regression | 2.12 | 4.90 | 2.21 | 0.90 |
| XGBoost Regression | 7.94 | 75.44 | 8.69 | -0.54 |
| Random Forest | 6.71 | 84.52 | 9.19 | -0.72 |
| Decision Tree Regressor | 7.82 | 75.34 | 8.60 | -0.54 |
| SVR | 25.67 | 683.26 | 26.14 | -1965.68 |

The Gradient Boosting Regressor revealed this sensitivity in terms of the past data through a sudden rise in its future predictions since 2019. Notably, predictions fall within a narrow 95% confidence interval signifying high specificities regarding model parameters. Nonetheless, an apparent case of overfitting may have led to the rather aggressive reaction of the model to recent data tendencies, a scenario where it has become extremely tuned on historical patterns (Figure 6a). Indeed, MAE = 2.15; MSE = 8.27; R² = 0.83 imply good performance by spite of moderate overfitting tendency.

In addition, the predictions made by linear regression are more consistent and support a more uniform model that accurately relates to the real-time data available for the period 2019 to 2022. Because of its simplicity, this model can maintain a general trend without being disturbed by minor movements and is therefore very useful when a linear trend in the underlying data is taken into account. It has been noted that wider confidence intervals imply greater uncertainty in predictions. However, these predictions were very close to the actual trends, which underlines their credibility for linear forecasts (figure 6b). In terms of accuracy and reliability against this dataset, linear regression remains the most accurate, with the lowest MAE (2.12) and MSE (4.90) values, and an R² of 0.90, making it an ideal candidate for future forecasts. However, XGBoost regression presented a different picture. Initially, predicted values drop slightly before remaining constant. XGBoost is renowned for its ability to handle complex data models such as non-linear relationships and interactions, but it doesn’t seem to perform well with this dataset. On the one hand, the predictions are quite convincing due to a relatively narrow confidence interval, but on the other hand, the actual trend varies considerably from what was predicted, as shown in figure 6c. The dissonance is evident thanks to poor performance statistics; an MAE of 7.94, an MSE of 75.44 and a negative R² (-0.54) suggest that it cannot adequately represent the data’s underlying system.

Similarly, the Random Forest model, which averages multiple decision trees to reduce overfitting and increase predictive accuracy, predicts a mild decrease followed by a stable forecast. However, this ensemble model shows considerable uncertainty in its long-term predictions, with broad confidence intervals as depicted in Figure 6d. The metrics, including a MAE of 6.71, MSE of 84.52, and a negative R² of -0.72, confirm that this model performs poorly on this dataset and is not able to generalize well to future trends.

The Decision Tree Regressor performs similarly to XGBoost, with predictions that diverge notably from the actual data, as seen in Figure 6e. While decision trees are typically strong in handling non-linear data, this particular configuration did not yield useful predictions. With a MAE of 7.82, MSE of 75.34, and a negative R² of -0.54, this model is also overfitting to the historical data, leading to poor generalization in future predictions. The relatively large confidence interval further underscores the model’s uncertainty.

Lastly, the Support Vector Regressor (SVR) model shows the poorest performance among all models. The nearly flat prediction line, as shown in Figure 6f, indicates that SVR has failed to capture any meaningful patterns in the data. The very wide confidence intervals and the extreme values for the error metrics, including a MAE of 25.67, MSE of 683.26, and a negative R² of - 1965.68, highlight the model’s severe overfitting and complete lack of predictive power. This model is clearly not suitable for this dataset.

In summary, the Linear Regression model’s success in closely approximating the actual data from 2019 to 2022 in this study illustrates its effectiveness in capturing the primary trend without the complexity introduced by more sophisticated models. This outcome supports the use of Linear Regression in applications where data exhibits a primarily linear trend, providing a robust forecast with straightforward interpretability.

### Predicting new CL cases over the next 10 years

The historical data depicted (2009-2022) shows a gradual increase in the number of cases until 2019, followed by a sharp rise through 2022. Utilizing this data, the Linear Regression model projects a continued linear increase in the number of new CL cases over the next decade (2023-2032), as illustrated by the red dotted line (**Fig 7**).

The predictive model’s 95% confidence interval, shown in gray, indicates the range within which future case numbers are expected to fall with 95% certainty, assuming the current trend continues without significant deviation. The width of the confidence interval suggests a reasonable degree of uncertainty in these predictions, reflecting potential variability in future trends based on the historical data pattern.

The results from the Linear Regression model show a relatively positive performance based on several key evaluation parameters. The MAE of 9.36 suggests that the model’s predictions, on average, deviate from the actual recorded data by approximately 9.36 cases. This level of MAE indicates a moderate level of error in the context of public health predictions, where precision can be critical for resource allocation and planning.

Furthermore, the MSE stands at 108.80, and the RMSE is calculated to be 10.43. These metrics, particularly the RMSE, provide a more sensitive measure of the model’s prediction errors by emphasizing larger errors more significantly due to the squaring of each difference. An RMSE of 10.43 is quite reasonable, suggesting that the model’s predictions are relatively close to the true data points, considering the scale of the case numbers involved.

Most notably, the R² value of 0.74 illustrates that approximately 74% of the variability in the number of new CL cases is explained by the model. This is a robust indicator of the model’s effectiveness, showing a strong linear relationship between the predictors used and the outcomes predicted. An R² of 0.74 in a public health context, especially for a disease like CL that may be influenced by numerous environmental and socio-economic factors, reflects a high degree of model reliability and utility.

Overall, these metrics underscore the Linear Regression model’s competency in forecasting CL cases, highlighting its potential as a tool for public health officials in anticipating disease trends and preparing accordingly. This model’s predictive accuracy, combined with its interpretability, makes it a valuable asset in the strategic planning and management of CL. However, continuous monitoring and model validation against new data will be essential to maintain the accuracy and relevancy of the predictions.

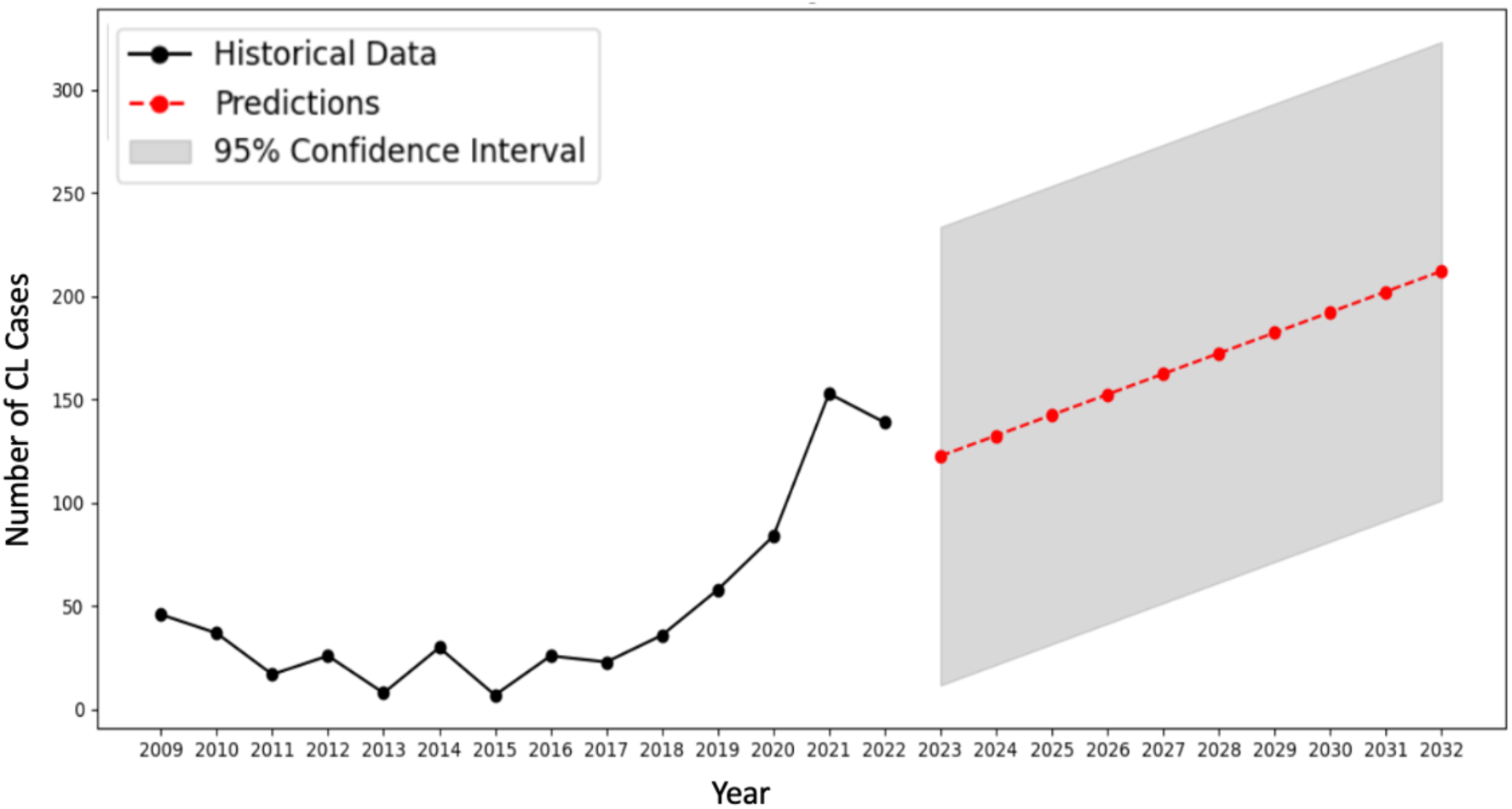

Prédiction des nouveaux cas de LC au cours des 10 prochaines années par modèle de régression linéaire (intervalle de confiance à 95 %).

## Discussion

Cutaneous leishmaniasis is still a major health problem in Morocco with more than 2,000 cases per year [19]. Therefore, a deep understanding of the disease epidemiology and distribution is crucial to implement targeted strategies. In this context, our study aims to decipher the epidemiology of CL in a region of major significance in Morocco. Casablanca-Settat region is home to more than 6 million residents and the location of Morocco’s largest city, Casablanca. To the best of our knowledge, this is the first study that analyzes spatio-temporal trends of CL in this Moroccan region. The region saw its first case of CL due *L. tropica* in 2006 and subsequently, 553 occurrences of CL between 2007 and 2012 were recorded, with the majority originating from one locality, El Brouj [14]. Notably, all recorded cases during this period in the region were attributed to *L. tropica.* From 2010 to 2012, health authorities initiated a response strategy with active case detection within communities in El Brouj to ensure early diagnosis and treatment and raise awareness amongst the population, thus leading to the decrease of cases and the observed low peak of 2015. These interventions, when implemented alongside other public health measures, can significantly contribute to the control and prevention of CL in endemic areas. Even though animal infections were recorded, *L. tropica* is still considered to be mainly anthroponotic in the country and could therefore spread more easily considering the numerous human travel movements observed in the Casablanca-Settat region, as long as *P. sergenti* is present [9].

Regarding clinical epidemiology data, we observed that disease incidence was generally higher in women than men throughout the whole study period, which is concordant with what has been previously reported by Amarir *et al* in 2015 in Settat province, who highlighted that this disparity could be due the lack of seeking medical care by men compared to women [14]. We also report a higher incidence in children between 5 and 14 years old, which is also in the line of their study, as children spend more time outdoors playing and are in closer contact to sandfly breeding sites.

The present study is a 14 years retrospective analysis of CL distribution in the region. We observed a steady increase in cases until 2019, followed by an exponential rise, reaching a peak in 2021. We also noted a seasonal variation of the number of recorded cases during the study period, as most cases were recorded during the spring from March to June, which was supported by the GAM analysis and is concordant with previous studies from Iran, Afghanistan and Spain that reported seasonal variations of CL cases (caused by various species), indicating that seasonal climate conditions may play a role in the disease cycle [20–22]. The increase in cases during spring could be attributed to behavioral and transmission factors, with a lag between the end of the transmission season (summer), the incubation period, lesion apparition and the time it takes for patients to seek medical care.

Seasonal variation is not the only factor contributing to the number of recorded cases as incubation period plays a critical role as well [23]: it can be defined as the interval of time between the parasite inoculation by the vector and the initial CL lesion. Estimating the incubation time would be useful for controlling CL since it would reveal the peak period of parasite transmission [24]. However, a research gap regarding incubation period estimation is observed in Morocco mainly because of the delay in seeking medical consultation, that could also influence CL incidence.

Cutaneous leishmaniasis cases in the study region are mainly distributed in certain localities, with one locality in particular standing out – El Brouj. The latter has consistently reported the highest number of CL cases over the entire period ’study. Given the status of El Brouj as an active focus, we examined the impact of climatic variables on the distribution of CL within this locality. By focusing our analysis on El Brouj, we aimed to better understand how climate might impact CL transmission, where the number of reported cases has been highest. Using a linear regression model, we analyzed the impact of climatic variables on the incidence of CL over 14 years (2009-2022). We showcase a strong positive correlation between both minimum temperature and precipitation and the incidence of CL, in agreement with Bounoua *et al* [25]. The increase in temperature has enabled sand fly larvae to survive the winter, fostering CL endemicity where none previously existed. Recently, a study demonstrated a significant association between both temperature and humidity and the incidence of CL due to *L. tropica* in Imintanout (South-West Morocco), highlighting that small elevations in temperature impact parasite life cycle within its vector [12]. Furthermore, another recent study identified increased rainfall as a critical factor in the proliferation and survival of the CL vector [26].

On the other hand, using CL cases data from 2009-2018 and climate variables (T° Max, T° Min, mean T°, precipitation and humidity), we tested and compared four machine learning models to predict CL cases in El Brouj for the years 2019-2022. We then observed that the Linear Regression model was the most successful in closely approximating the actual data from 2019 to 2022. This illustrates its effectiveness in capturing the primary trend without the complexity introduced by more sophisticated models. This outcome supports the use of Linear Regression in applications where data exhibits a primarily linear trend, providing a robust forecast with straightforward interpretability.

Thus, we performed a prediction analysis using Linear Regression model for the next 10 years of CL cases in El Brouj. The model shows an increased projection of future CL cases accordingly with present data. Overall, these metrics underscore the Linear Regression model’s competency in forecasting CL cases, highlighting its potential as a tool for public health officials in anticipating disease trends and preparing accordingly. This model’s predictive accuracy, combined with its interpretability, makes it a valuable asset in the strategic planning and management of CL. However, continuous monitoring and model validation against new data will be essential to maintain the accuracy and relevancy of the predictions. We believe that vector presence data in the region with future climate data forecast could allow the model to better predict future trends of CL in the region. While the short period for analyzing grouped spatial data and the inherent limitations present challenges, a deeper understanding of the historical dynamics CL in the studied area offers hope that, with more comprehensive data and advanced modeling techniques, future predictions can be refined to better inform effective interventions. Moreover, climate change should also be taken into account since it could also heavily influence future forecasts, as extreme temperatures or heavy rainfall can impact multifactorial diseases such as CL.

Since 2008, it has been pointed out that the distribution of CL is impacted by climate change, in particular temperature variations, which may directly affect the abundance of sand flies or on the parasite growth process within a vector [28]. Consequently, temperature and precipitation are two restrictive factors that determine the extent of vector activity, and the onset and evolution of CL [29]. A 2023 ecological niche modeling study indicates that *P. sergenti* will likely spread into regions that are now deemed unsuitable for the species. For the following 30 years, *L. tropica* is anticipated to have a significant expansion in Morocco’s southern regions [30]. Devastating effects of climate change are particularly felt by emerging nations. Future climate forecasts indicate a strong probability of less precipitation and elevated temperatures across most of Africa including Morocco, which might worsen its future, especially in terms of climate sensitive infectious diseases spread [31].

In addition to climatic and environmental factors, other causes could be contributing to the noticed increase in the number of CL cases in Casablanca-Settat region, especially socio-economic and human factors. First and foremost, the lack or deficient awareness of the disease cycle and symptoms by both population and local healthcare professionals hinders the efforts of preventive measures taken by the authorities. Moreover, knowledge gaps differ among community members, highlighting the need for targeted health promotion and education tailored to the specific roles and perceptions within the community.

Another factor thought to increase the incidence of CL is urban sprawl. The area is fast becoming more urbanized as a result of the building of new residential complexes in places with a high concentration of sand fly breeding grounds/sites, as well as roads and highways making human movements easier, and also irrigation dams. Due to its strategic location as an unavoidable route between the South and the North as well as its industrialization, the Casablanca-Settat region became an administrative entity in the early 20^th^ century [14]. As Casablanca is the economic capital of the country and the land of all opportunities, people migrate from all over the country to seek jobs and they might come from CL endemic regions. According to a recent study carried out at the Ibn Rochd University Hospital in Casablanca, 75% of patients with CL had visited endemic areas of Morocco, highlighting the risk of infection when visiting such areas. Additionally, patients with *L. tropica* were reportedly infected across 9 different regions, reflecting the species’ propensity to spread geographically into previously non-endemic areas. As revealed by Ait Kbaich *et al*. (2017) and El Alem *et al*. (2018), a number of studies have demonstrated the spread of *L. tropica* to diverse regions, including those formerly afflicted by *L. major* or *L. infantum*, suggesting shifting geographical trends [32–34]. The 2018 study also highlights the connection between increased risk of CL due to *L. tropica* and socioeconomic factors in southwestern Morocco [34]. In addition, the increased risk of human exposure to sand flies could be due to the placement of houses in low income settings, which are frequently found in congested locations with poor hygiene and close to waste disposal sites. The spread of leishmaniasis can be accelerated by inadequate public hygiene measures such as the lack waste management and inappropriate sanitation facilities, which can also serve as sandfly breeding grounds/sites thus promoting human-vector contact. Living in regions where residents have insufficient health care facilities, is another risk factor [35]. Presently, the seaside of the region has still been indemn of any CL cases during our study time period. However, increased urbanization together with climate change and population movements could lead to the introduction of CL due to *L. tropica* in these areas, as *P. sergenti* is a flexible species, with a broader range of optimal climatic conditions compared to the vectors of *L. major* and *L. infantum*, and its presence is expected to reach all the intact provinces of Casablanca-Settat region according to a prediction model [30].

Finally, the cases included in this study are the ones notified to the Ministry of Health and Social Protection. However, underreporting of CL is still an issue in our country. Patients with small lesions may not always be sent to health centers, which could impact the number of cases that are identified and recorded in some locations. In contrast, the increase in cases observed after 2019 could be due to better disease knowledge and better patient care. Therefore, prevention through the reduction of contact between humans and vectors and decreasing vector population are key to its prevention. Some key measures to achieve this reduction include active screening, early diagnosing and treatment and covering the lesions to avoid further vector blood meals. In Morocco, despite an active national program of fight against leishmaniases, surveillance is mainly limited to active foci which may lead to under-reporting of CL cases [8]. Thus, the rise in cases in the Casablanca-Settat region highlights the need to reinforce and sustain local programs, but also better better monitor the disease spread and improve case notification.

## Conclusion

Considering the endemic situation of CL in Morocco, further research is necessary to map the presence of the disease throughout the country, especially in regions where it has not been extensively studied. This study is a first step towards the documenting and surveilling epidemiological trends in the area and highlighting gaps that need to be addressed in terms of case prediction.

The rise in CL cases shows the necessity to strengthen control and surveillance measures specifically tailored to the Casablanca-Settat context, especially the Settat province, and further study and map the risk in the other provinces of the region.

Targeted programs must be re-inforced when outbreaks are occurring on a local level. This will involve working closely with healthcare facilities to promote accurate and timely reporting of new infections. It also means conducting field investigations to map hotspots, raise awareness among both the population and physicians, and identify favorable environmental risk factors. Finally, a good monitoring of the spread of CL through an integrated surveillance system is essential to establish an effective public health response. Strengthening collaboration between public health authorities, research institutions, and local communities is crucial for implementing sustainable control and surveillance measures. A coordinated approach specific to the region would make it possible to control any future increase in CL cases in the Settat province, thus reducing its incidence in this densely populated region.

## Data Availability

All data produced in the present study are available upon reasonable request to the authors

## Acknowledgments

The authors are grateful to Professor Aboubaker Farah for his assistance with QGIS mapping. This study was carried out in the frame of a research project on cutaneous leishmaniasis in Morocco funded by the Moroccan Ministry of National Education, Professional Training, Higher Education and Scientific Research (CNRST, PPR/2015/27).

## Author contributions

IE: Conceptualization, Methodology, Writing – original draft. HT: Conceptualization, Methodology, Writing – original draft. BB: Data Acquisition, Validation, Editing. SB: Data Acquisition, Validation, Editing. RF: Review & editing. MR: Conceptualization, Methodology, Supervision, Validation, Writing – review & editing. ML: Conceptualization, Methodology, Supervision, Validation, Writing – review & editing.

